# Depression Incidence in Patients with Hypertension in a Single Outpatient Center

**DOI:** 10.1101/2024.09.24.24314333

**Authors:** Richard Amoateng, Natthew Arunthamakun, Xiarepati Tierliwaerdi, Alexandra Johnston, Mrudula Gadani

## Abstract

**Introduction:** Hypertension and depression are both highly prevalent in the United States and each condition is commonly managed in the outpatient setting. This study aims to investigate the incidence of depression in patients who are diagnosed with hypertension.

**Methods:** A retrospective cohort study was conducted analyzing 9,240 adult patients at a single center outpatient clinic between 2019 and 2022. Blood pressure and PHQ-9 from the latest outpatient visit were recorded. Demographic data associated with hypertension were also obtained. Bivariate statistical analysis was performed. Multivariate linear and logistic regression models were adjusted for co-morbid conditions.

**Results:** Out of the 9,240 patients studied, 3,694 (40%) had a diagnosis of hypertension. Patients with hypertension were more likely to be older (61.96 ± 14.95 vs 39.85 ± 15.61, p=0.000), male (37% vs 33%, p =0.002) and black (45.2% vs 23.3%, p<0.001) compared with those without a diagnosis of hypertension. The mean PHQ-9 score was higher in patients with hypertension than in those without (2.97 ±4.66 vs 2.70 ± 4.93, p=0.000). Patients with hypertension were more likely to have uncontrolled depression defined as PHQ-9 score >4 (22.55% vs 19.4%, p<0.001) even when adjusted for co-morbid conditions (adj OR 1.216 95% CI 1.06 -1.34 p=0.005).

**Conclusions:** In this cohort of patients, a diagnosis of hypertension was associated with an increased rate of uncontrolled depressive symptoms. Hence patients with hypertension should be screened using validated PHQ-9 tools in the outpatient setting and offered appropriate treatment for depression.

## Introduction

Hypertension (HTN) is a significant public health problem and patients are routinely screened for this condition in the outpatient setting. Over 116 million patients, representing 47% of all adults in the U.S., suffer from hypertension[1]. Patients with hypertension face increased risks of stroke, cardiovascular disease, premature disability, and even death [2, 3]. These negative outcomes related to hypertension are associated with significant psychosocial stress to patients, making them more susceptible to developing psychological disorders.

Depression is a common psychological disorder associated with poor health outcomes and comorbid conditions. According to the 2020 National Survey of Drug Use and Health (NSDUH), an estimated 21.0 million U.S. adults aged 18 or older had at least one major depressive episode [4]. It is well established that depression is a common risk factor for multiple conditions such as cardiovascular disease [5, 6], but depression most often goes undiagnosed which contributes to ineffective control of the primary disease.

Hypertension and depression are both highly prevalent in the United States. Several clinical trials have highlighted the prevalence of depression in patients with hypertension [7-10]; however, most of these studies were conducted in China. Further studies are needed to better define the relationships between these conditions. Accordingly, this study aims to investigate the incidence of depression in patients who are diagnosed with uncontrolled hypertension in the outpatient setting.

## Methods

All patients at single outpatient primary care center were retrospectively reviewed. Patient demographics were obtained from electronic medical records. The study was approved by the Institutional Review Board at Allegheny Health Network.

Records of patients with prior diagnosis of hypertension, and comorbidities were obtained. Blood pressure measurements were obtained at each visit.

HTN was defined according to the 2017 AHA/ACC hypertension guidelines. Stage I HTN was defined as a systolic blood pressure >130 mmHg or a diastolic blood pressure >80 mmHg. Stage II HTN was defined as a systolic blood pressure >140 mmHg or diastolic blood pressure >90mmHg. [3] For the purposes of this study, all Stage II HTN was defined as uncontrolled hypertension.

Patients were screened for depression using the nine-item patient health questionnaire-9 (PHQ-9) tool. The PHQ-9 screening tool contains items derived from the DSM-IV classification for major depression disorder and pertains to: (1) anhedonia, (2) depressed mood, (3) insomnia, (4) fatigue, (5) appetite change, (6) guilty feeling, (7) concentration difficulties, (8) restless feeling or feeling down and lastly (9) suicidal ideations. Patient answered each question on a 4-point scale with a response of 0-3 based on frequency of their symptoms: (0) no symptoms; (1) several days; (2) more than half the days; or (3) nearly every day.

### Statistical Analysis

All data were analyzed using Statistical Package for the Social Sciences (SPSS) version 28 for Mac. Bivariate analyses were performed. Student’s t-test was used for quantitative variables such as age. Chi-square and Fischer-Exact test were used for categorial variables, hypertension diagnosis, PHQ-9 score, and absence or presence of comorbidities. A multivariate regression analysis was performed. Adjusted Odds ratio and corresponding 95% confidence intervals were obtained. A p-value <0.05 was considered statistically significant. Data are expressed as mean ± standard deviation.

## Results

### Baseline characteristics

Demographic characteristics of 9,240 patients are outlined in Table 1. A total of 3694 (40%) patients carried a diagnosis of hypertension. Most of the study population were female; however, a diagnosis of hypertension was most strongly associated with male sex (42% of males had hypertension compared to 39% of females, p <0.002). The study population was predominantly White (n=5566, 60%). A total of 1898 (34%) White patients had a chart diagnosis of hypertension compared to 1671 (56%) of 2960 Black patients (p<0.001).

**Table 1.**
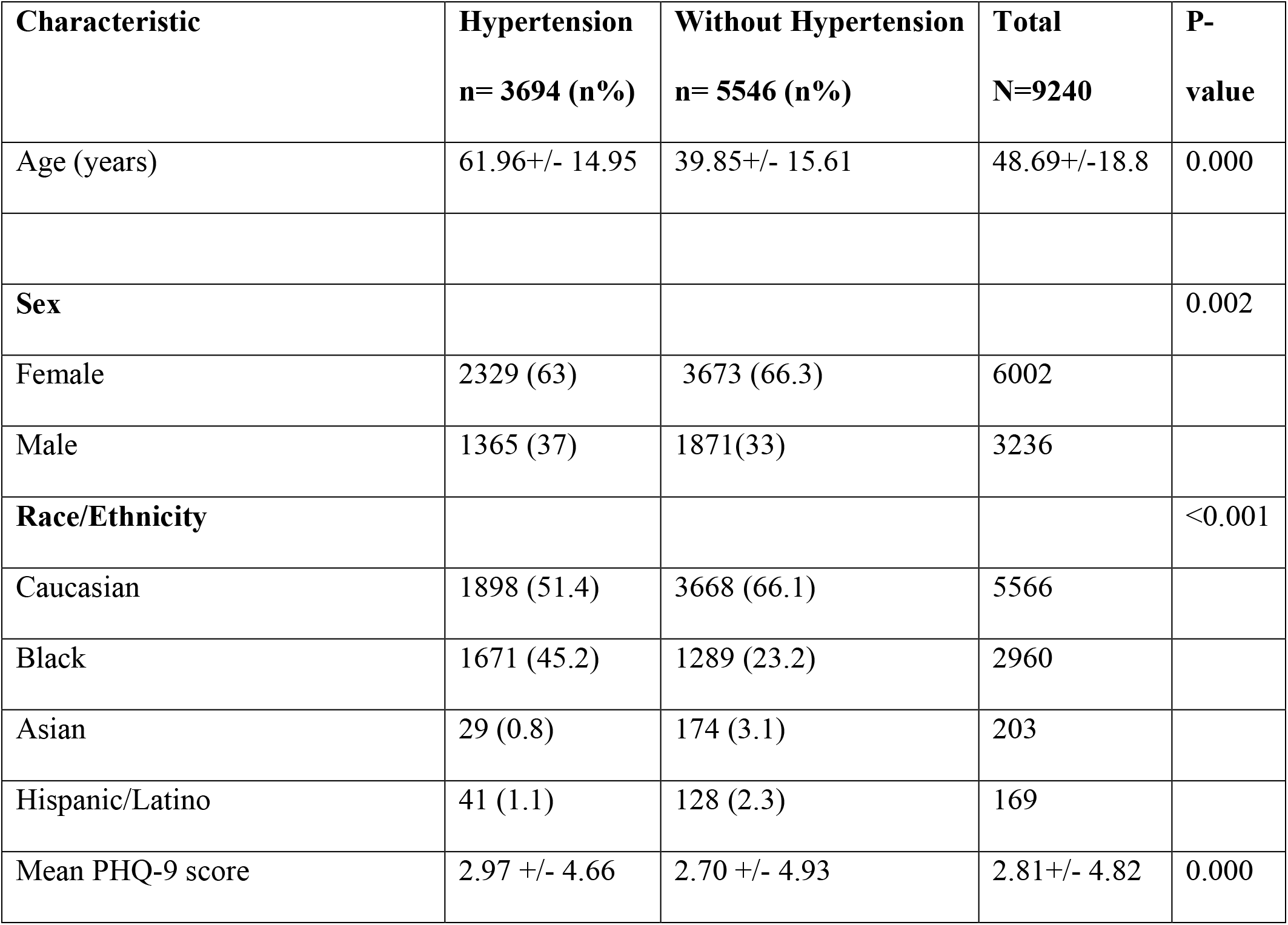
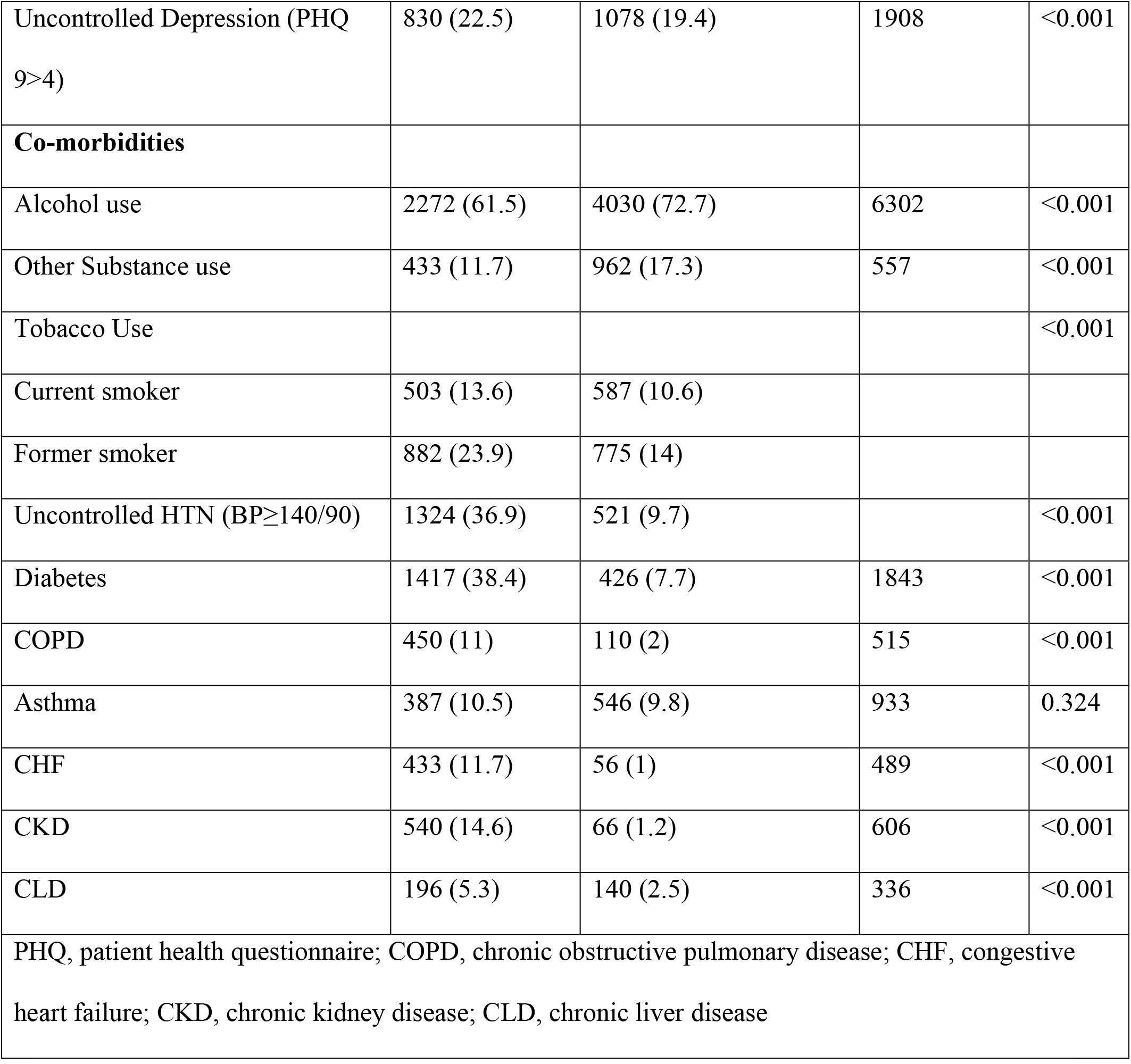
Baseline characteristics.

### Depression and Hypertension

In the entire cohort, 7296 (79%) had controlled depression or were not depressed as defined by PHQ-9 <5. The mean PHQ-9 score for patients with hypertension was higher than for patients without hypertension (2.97± 4.66 vs 2.70 ± 4.93, p=0.000). Of all patients with hypertension, 842 (23%)) had uncontrolled depression and PHQ-9 scores >4, as compared to 1097 (20%) in the non-hypertensive group. Mild depressive symptoms (PHQ-9 5-9) were most strongly associated with hypertensive diagnosis (17.5% vs 12.8%, p <0.001). In addition, 245 patients had severe depression, 95 (2.5%) of whom were in the hypertensive group (Table 2)

**Table 2.** Depression severity by PHQ-9 in hypertensive patients.

| Depression severity by PHQ-9 | Hypertension | Without Hypertension | Total |
| --- | --- | --- | --- |

|  | **n= 3694 (n%)** | **n= 5546 (n%)** | **N=9240** |
| --- | --- | --- | --- |
| 0-4 (Not depressed) | 2848 (77.2) | 4448 (80.2) | 7296 |
| 5-9 (mild depression) | 646 (17.5) | 711 (12.8) | 1357 |
| 10-14 (moderate depression) | 61 (0.7) | 130 (1.4) | 191 |
| 15-19 (moderately severe depression) | 44 (1.2) | 103 (1.9) | 147 |
| >19 (severe depression) | 91 (2.5) | 153 (2.8) | 245 |
| PHQ, patient health questionnaire |  |  |  |

### Comorbidities

Traditional cardiovascular risk factors were common in the population studied (Table 1). In the hypertensive group, 1324 (37%) patients had ACC/AHA stage II hypertension which was classified as uncontrolled hypertension in this study. Diabetes, chronic kidney disease (CKD), tobacco use, chronic obstructive pulmonary disease (COPD), and congestive heart failure (CHF) correlated most strongly with hypertension (P-values <0.001). Interestingly, alcohol use and other illicit substance use showed a lesser association in the hypertensive group compared to patients without hypertension (62% vs 73% and 12% vs 17% respectively, p-value <0.001).

### Multivariate regression analysis ((Table 3)

Using a multivariate regression analysis, uncontrolled depression (PHQ-9> 4) was independently associated with hypertension diagnosis. Hypertensive patients were 1.2 times more likely to have uncontrolled depression than patients without the condition (1.216 OR 95% CI 1.06-1.40, p <0.05). Among other comorbidities, diabetes was most strongly associated with hypertension, followed by congestive heart failure, chronic kidney disease (3.414 OR 95% CI 2.96-3.93, p<0.001; 2.912 OR 95% CI 2.10-4.03, P<0.001; 2.633 OR 95% CI 1.95-3.55, p<0.001; 2.379 OR 95% CI 2.11-2.69, respectively). Black race was also associated with hypertension in the population studied (p <0.001).

**Table 3.** Multivariate regression analysis of all variables on hypertension diagnosis.

| **Variable** | **OR** | **95% Confidence Interval** | **P-value** |
| --- | --- | --- | --- |

|  |  | **Lower** | **Upper** |  |
| --- | --- | --- | --- | --- |
| Uncontrolled<br>Depression:<br>PHQ-9>4 | 1.216 | 1.059 | 1.395 | 0.005 |
| Age | 1.076 | 1.071 | 1.080 | <0.001 |
| Black Race | 2.379 | 2.105 | 2.687 | <0.001 |
| Asian Race | 0.534 | 0.329 | 0.867 | 0.011 |
| Female Sex | 0.668 | 0.594 | 0.752 | <0.001 |
| Tobacco Use | 1.081 | 0.911 | 1.281 | 0.372 |
| Alcohol Use | 1.295 | 1.142 | 1.468 | <0.001 |
| Other illicit<br>drug use | 0.929 | 0.714 | 1.187 | 0.522 |
| CHF | 2.912 | 2.102 | 4.034 | <0.001 |
| Diabetes | 3.414 | 2.964 | 3.931 | <0.001 |
| COPD | 1.357 | 1.051 | 1.753 | 0.019 |
| Asthma | 1.106 | 0.924 | 1.323 | 0.273 |
| CKD | 2.633 | 1.954 | 3.549 | <0.001 |
| CLD | 1.1 | 0.831 | 1.456 | 0.505 |
| PHQ, patient health questionnaire; COPD, chronic obstructive pulmonary disease; CHF, congestive heart failure; CKD, chronic kidney disease; CLD, chronic liver disease |  |  |  |  |

## Discussion

In our single-center outpatient setting, nearly 40% (n=3,694) of 9,240 patients had a diagnosis of hypertension, and of those, almost half (1,324) were classified as having uncontrolled hypertension with blood pressure ≥140/90. These individuals scored higher on PHQ-9 screening with a greater likelihood of uncontrolled depression compared to those in the non-hypertensive group. This was even true after adjusting for other comorbid conditions. Other factors that placed individuals at risk for uncontrolled depression in both groups included increased age, male sex, and Black race. Overall, our analysis adds support to existing evidence that patients with hypertension are more likely to develop depression and should be appropriately screened using validated tools in the outpatient setting.

In the present study, multivariate regression analysis revealed that patients with concomitant comorbid conditions were more likely to have hypertension, the cardiovascular outcomes of which have been well-described in the literature [11]. While being of Black race showed a 2.38 odds ratio of developing hypertension, Asian race appeared to be least associated with hypertension diagnosis with an odds ratio of 0.53. Further subgroup analysis revealed that Black patients were more likely to have experienced health disparities and were disproportionately affected by health inequities. These trends may contribute to the greater likelihood of developing uncontrolled depression and uncontrolled hypertension among Black patients [12, 13]. The seemingly lower odds of hypertension in the Asian population seen in the current analysis requires further study.

Individuals with chronic diseases such as hypertension may be at increased risk of depression [9, 14]. A systematic review and meta-analysis of observational studies by Li et al. showed that in 41 studies with a total of 30,796 individuals, one-third of hypertensive patients had depressive symptoms when screened by clinical interview [9]. However, a cohort study by Obas et al. found that in normotensive people, the age-related increase in systolic BP was lower by about 2 mmHg among patients with depression compared to participants without depression over a 10-year time frame [15]. It was also concluded that the presence of depression at baseline increased the odds of hypertension diagnosis. Similarly, a study of ethnic minority groups in the Netherlands showed no associations between significant depressed mood and hypertension management [16]. Despite these mixed results in the literature on the relationship between hypertension with depression, there is a growing consensus that the two conditions share an association. Screening for depression in individuals with hypertension (and other chronic comorbidities) should be performed during outpatient visits, as addressing mental illness among chronically ill individuals is necessary to improve quality of life [17].

Our results successfully demonstrated the usefulness of the validated PHQ-9 screening tool, which should be widely incorporated in most patient encounters in the clinical setting. Screening for depression is conducted routinely in primary care clinics via patient-reported questionnaires. The most widely used tool is the Patient Health Questinnaire-9 (PHQ-9), which has a sensitivity of 74% and a specificity of 91%. [17] The tool consists of 9 items, each of which is scored on ascale of 0-3. The questionnaire is therefore scored between 0-27, with scores above 10 indicating a likely depressive disorder. As part of a two-step approach, a short preliminary questionnaire called the Patient Health Questionnaire-2 (PHQ-2) may be used before PHQ-9 as pre-screening, with positive screening warranting the more detailed PHQ-9. The use of PHQ-2 has been shown to decrease the need for PHQ-9 screening, but sensitivity and specificity are almost the same whether the tools are used alone or together [18].

## Strengths and Limitations

Major strengths of this study include a large sample size which was statically powered and inclusion of a diverse patient population. The cross-sectional designs limit the cause-effect relationship between hypertension control and depression. The study was also limited by the fact that medication therapies were not obtained.

## Conclusion

In conclusion, a diagnosis of hypertension was associated with an increased rate of uncontrolled depressive symptoms. Black race and co-morbid conditions like diabetes and congestive heart failure were also significantly associated with hypertension. Hypertensive patients should be appropriately screened using validated PHQ-9 tools in the outpatient setting and offered appropriate treatment for depression.

## Data Availability

All data are available upon request

## Abbreviations

CHF: ongestive heart failure
CLD: chronic liver disease
CKD: chronic kidney disease
COPD: chronic obstructive pulmonary disease
HTN: hypertension
PHQ: patient health questionnaire

## Acknowledgments

RA performed the statistical analysis, edited, and wrote portions of the manuscript. NA wrote the bulk of the discussion. XT wrote the introduction. AJ and MD edited and reviewed the manuscript.

